# LLM-Based Classification of Case Report Abstracts: A Pilot Study on Interactions between Radiotherapy and Systemic Therapies

**DOI:** 10.64898/2025.12.22.25342797

**Authors:** Fabio Dennstädt, Til Bobnar, Alin Handra, Paul Martin Putora, Irina Filchenko, Sarah Brüningk, Daniel M Aebersold, Nikola Cihoric, Mohamed Shelan

## Abstract

**Background:** The growing volume of biomedical literature, especially in oncology, necessitates automated tools for extracting clinically relevant information. Large Language Models (LLMs) offer promising capabilities for data extraction in this domain. However, their potential to extract clinically relevant information from case reports detailing rare treatment interactions, remains underexplored.

**Methods:** We systematically searched PubMed for case reports on interactions between radiotherapy (RT) and Pembrolizumab, Cetuximab, or Cisplatin. A random sample of 100 report abstracts for each therapy was manually classified by two independent medical experts using 17 Boolean questions about patient demographics, treatment, cancer type and outcome with mutually exclusive answers, forming a ground truth. An LLM-based system with the open-source GPT models (GPT-OSS-120B and GPT-OSS-20B) was applied to classify these reports and the remaining dataset entries using the defined question structure. Performance of the LLM-based information extraction was evaluated using the standard classification metrics accuracy, precision, recall, and F1-scores.

**Results:** The systematic searches yielded 320 (Pembrolizumab), 147 (Cetuximab), and 2055 (Cisplatin) publications. Inter-rater agreement for manual classification was high (Cohen’s kappa = 0.87), though lower (0.60-0.80) for specific outcome and cancer type questions. The LLM-based classification (GPT-OSS-120B model) achieved high overall performance with an F1-score of 94.33% (95.83% accuracy, 93.69% precision, 94.98% recall). Performance was consistent across systemic therapies, with the smaller GPT-OSS-20B model showing similar results (F1-score 94.06%). Analysis of the entire datasets revealed that 56.02% of publications described patients who received both RT and systemic therapy. Proportions of positive and negative outcomes varied by therapy and sequencing.

**Conclusions:** LLM-based classification systems demonstrate high accuracy and reliability for curating scientific case reports on RT and systemic therapy interactions. These findings support their potential for high-throughput hypothesis generation and knowledge base construction in oncology, particularly for underutilized case reports, with even smaller open-source models proving effective for such tasks.

## Introduction

The increasing number of biomedical publications, particularly in oncology, has created an urgent need for automated tools to extract clinically relevant knowledge from unstructured text.

Generative Artificial Intelligence (AI) and Large Language Models (LLMs) are transforming the landscape of automated text analysis and natural language processing (NLP) in the biomedical literature (1). LLMs can synthesize evidence, answer clinical questions, and generate summaries from unstructured data (2), (3), (4).

LLMs are increasingly being used in citation screening (3), in scoping reviews (5) and systematic literature reviews (6) as well as for data extraction from the publications or trial protocols (7). In one of our own studies, we observed high levels of performance with F1-scores >92% when using LLMs to retrieve clearly defined information from oncological trials (8). Nevertheless, critical gaps persist, and it is still unclear how broadly and in which context LLMs can successfully be used to assist in knowledge generation and analysis of biomedical literature.

Case reports, which detail rare events, novel therapeutic combinations, and unexpected adverse interactions, are a rich but underutilized resource to learn from clinical experience and generate hypotheses (9) (10). Manual classification and comparison of case reports is labor-intensive, time-consuming, and subject to bias and inconsistency (11). This is especially true for complex clinical questions that require domain knowledge. While case reports describe unique situations and cannot be used to deduce causality about an outcome, they are invaluable for generating hypotheses in rare situations. They can furthermore be used to obtain knowledge about interactions of individual factors and therapies.

In this work, we aimed to investigate if LLMs could be used for basic classification of case reports on interactions between radiotherapy (RT) and different systemic therapies. Combining these treatments can result in favorable outcomes such as an increased therapeutic effect (12) in some situations, but can lead to severe and rare side effects in other situations (13). Such interactions are still largely unknown, but highly relevant in clinical practice as physicians often do not know when a combination of RT with a specific systemic therapy is safe. This is true for widely used substance of different therapies, like the immunotherapy Pembrolizumab, the targeted therapy Cetuximab, and the chemotherapy Cisplatin. Despite being used in clinical routine, detailed interaction data of such substances with RT in rare clinical scenarios remains limited.

Automatic analysis of case reports on such interactions could be highly valuable and assist in overall knowledge synthesis, as well as in the generation of guidelines or consensus recommendations (14). Interactions between RT and systemic therapies are complex, with diverse terminology, ambiguous reporting, and subtle temporal relationships that vary across different cancer types. Traditional NLP methods are often limited in their ability to analyze these nuances at scale. Using an LLM-based approach that can conduct an analysis across different cancer types and oncological scenarios, could enabling large-scale, automated knowledge synthesis. This facilitates the identification of emerging patterns of toxicities and synergies from the collective evidence in case reports, revealing insights that are not apparent from isolated clinical cases.

Therefore, this study aimed to investigate whether an LLM-based system can automatically classify scientific case reports on RT-systemic therapy interactions with a level of performance comparable to human medical experts. We present the first systematic evaluation of such a system on the three systemic therapies Pembrolizumab, Cetuximab, and Cisplatin. These are highly relevant examples of an immunotherapy, a targeted therapy, and a conventional chemotherapy, all of which are commonly used in combination with radiotherapy for the treatment of various cancers.

## Methods

### Data Source and Search Strategy

We accessed PubMed (15) on January 10^th^, 2025, and conducted systematic searches for publications indexed as ‘Case Reports’ using structured search queries. Three queries were constructed to identify case reports involving RT and the systemic therapy Pembrolizumab, Cetuximab or Cisplatin, respectively. Search queries combined MeSH terms and therapy synonyms to cover a broad and comprehensive range of relevant literature (see **Supplementary File 1** for details and full queries). We obtained the titles and abstracts of the identified publications to create three datasets stratified by systemic therapy.

### Classification Protocol

We defined questions for basic, top-level classification of the case reports. The aim was to create classification questions with clear, mutually exclusive permissible answer options. We therefore created seventeen Boolean questions covering patient demographics, treatment, cancer type and outcome, each with the permissible answer options “TRUE” and “FALSE”. This allows to minimize ambiguities and enables determining the classification metrics recall, precision, and F1 for individual questions as well as for the entire datasets.

A basic structure logic of the questions with Boolean conditions was created, defining which questions are relevant in which context. For example, questions on therapy sequencing like “*Was Pembrolizumab given before Radiotherapy?”* were only asked as a follow-up question if both the questions “*Did the patient receive the systemic therapy Pembrolizumab?”* and “*Did the patient receive radiation therapy?”* were answered with “TRUE”.

Case reports that report only on the medical history of a single patient, usually provide clear information (already in the title and abstract), explaining the basic patient history. This allows for obtaining meaningful information and classifying the case reports into clinically relevant classifications. A basic overview of the approach with an example case report is provided in **Figure 1**. The case report was published by Lee et al. (16), reporting on a patient who received RT and Pembrolizumab and had an abscopal effect with complete disease remission.

**Figure 1:**
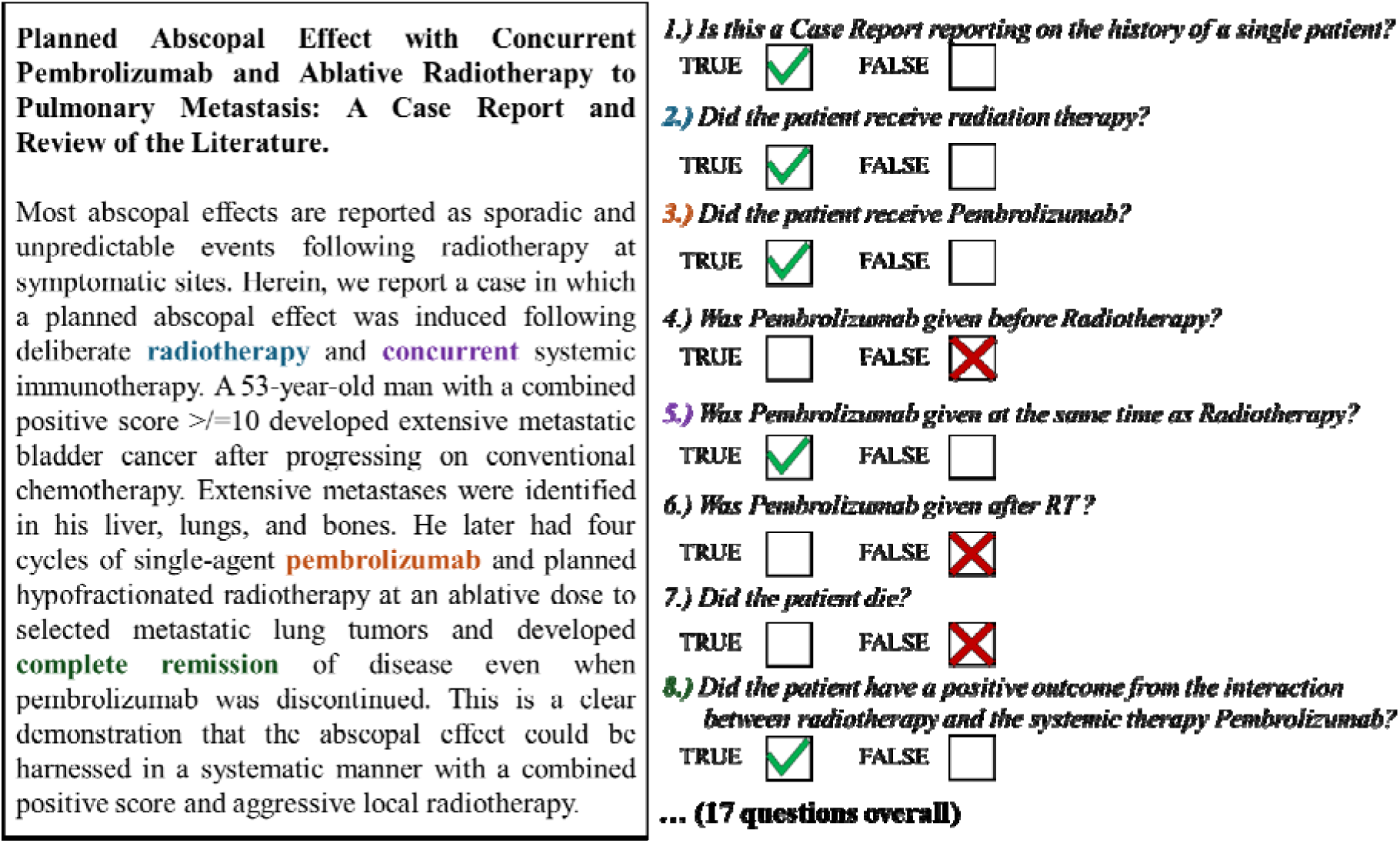
Basic overview and example on how the LLM-based classification of case reports works. Example case report published by Lee et al. (16). 17 Boolean questions were asked for each case report.

The first question to be asked was “*Is this a Case Report reporting on the history of a single patient?*”. This question is relevant to correctly identify publications that report only on the case of a single patient, since many of the publications indexed as “Case Report” on PubMed are case series or report about several patient situations.

Two follow-up questions were about whether the patient had received RT and the respective systemic therapy (Pembrolizumab, Cetuximab or Cisplatin). Additional follow-up questions addressed therapy sequence, vital status, interaction outcomes, patient gender and cancer type.

A list of all the individual questions used in the study is provided in **Table 1**. It should be noted that even related questions are not necessarily mutually exclusive. For instance, the questions regarding cancer type (Questions 12-17) are not exclusive, as a patient may have more than one cancer disease. Similarly, questions about therapy sequence (Questions 4-6) are not mutually exclusive, since radiotherapy and systemic therapy can be administered both concurrently and, in an adjuvant, or neoadjuvant setting.

**Table 1:**
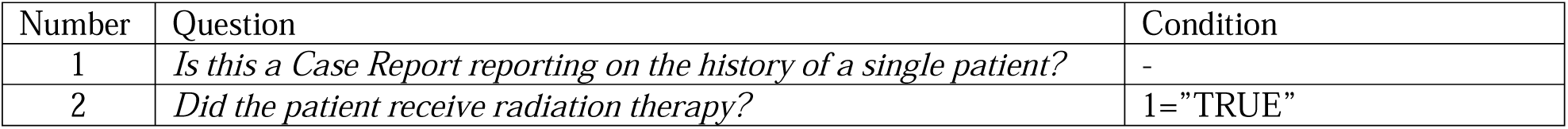
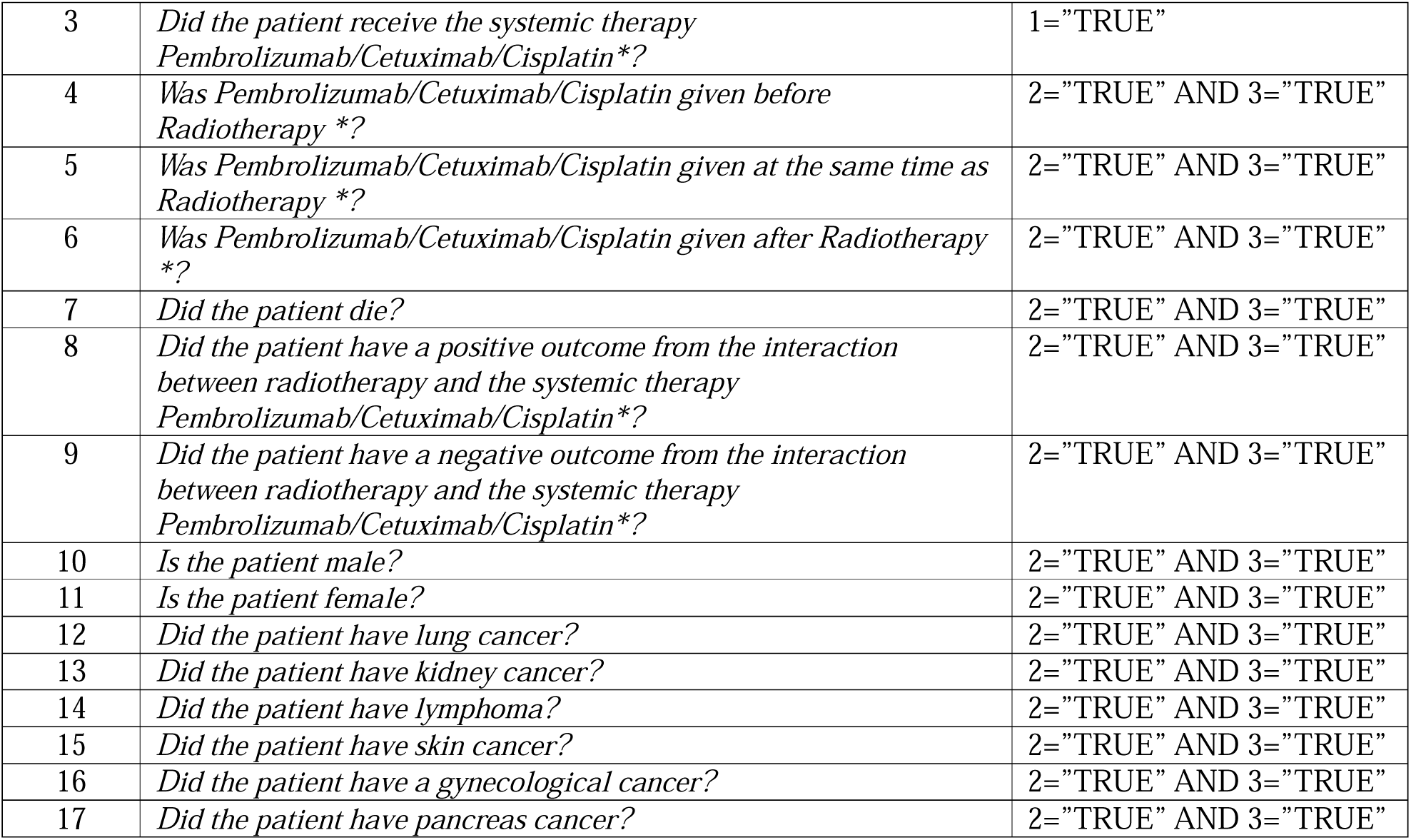
The classification questions used in the experiments. *Only the relevant systemic therapy was part of the question in each dataset.

Two questions (Questions 8 and 9) addressed “positive” or “negative” outcomes from the therapy interaction. These are also not mutually exclusive, as a patient could experience both a severe side effect and a good treatment response. In this case we intentionally did not provide strict definitions for these terms to avoid prematurely excluding relevant outcomes. Therefore, “positive” and “negative” were interpreted broadly to cover any interaction outcome described as beneficial or harmful.

### Manual Annotation and Ground Truth

For each of the three datasets (Pembrolizumab, Cetuximab, and Cisplatin) a random sample of 100 case reports was manually classified to obtain a ground truth for performance evaluation. This manual classification of each report was independently done by two researchers with medical domain knowledge (one radiation oncology resident with one year of clinical experience and one 6th year medical student). Cases of disagreement were resolved by the study coordinators. The ground truth datasets were used to benchmark LLM performance.

### Technical implementation of LLM-based extraction

The defined questions with the classification structure were implemented in a machine-readable .JSON format. The LLM-based classification was done with the *data-element-extractor* (v0.2.0) (17) library using the .JSON file and the default classification prompt of the library. This Python package provides an automated framework, which allows direct use of an LLM for a classification task by answering the questions (in the .JSON file) to a given text input.

The models GPT-OSS-120B and GPT-OSS-20B (18) were used for the classification in two independent evaluation runs. These LLMs originate from the same model family as the widely used ChatGPT models (e.g., GPT-4 or GPT5), but have been published open-source. We decided to use open-source models instead of the proprietary GPT models, as this ensures accessibility, transparency, and control of the models, preferable for application of generative AI in biomedical literature analysis (19).

Model inference was performed via the Cloud Service DeepInfra (20). No prompt engineering, fine-tuning or any other kind of optimization was applied. The default prompt of the Python package for classification tasks was used for all classification questions. The full data and code including the .JSON file for the structure together with further technical details are provided on GitHub and can be used to reproduce the results (21). The data extraction was performed on the ground truth dataset of 3×100 manually classified publications for the performance evaluation and in a second run on the remaining entries in the three datasets.

### Evaluation and Statistical Analysis

The study followed TRIPOD-LLM reporting guidelines (22) (**see Supplementary File 2**).

Statistical analyses were conducted in R version 4.0.1 (cran.r-project.org/). Continuous variables were presented as median and interquartile range, while categorical variables were presented as count (% of total). All statistical tests were two-sided and conducted at the 5% significance level. There was no missing data.

First, we calculated the Inter-rater Agreement (IRA) of the manual classification, which included 100 cases. This was done for each question, for the total set of questions per medication (i.e., Pembrolizumab, Cetuximab, and Cisplatin), and for the whole dataset (i.e., including the data of three medications) using Cohen’s kappa (23).

Second, we extracted the data on the entire dataset of cases using the LLM.

Third, we quantified the LLM performance (i.e., with accuracy, precision, recall, and F1-score) based on the 100 cases used for the ground truth dataset (see above). We quantified the Spearman’s correlation between IRA and F1-score. We also compared the LLM performance across 17 questions between three medications using pairwise Wilcoxon Signed-Rank Test with false-discovery rate correction for multiple comparison.

Fourth, we investigated the associations between the outcomes of the therapy (Questions 8 and 9) with therapy sequence (Questions 4-6), patient gender (Questions 10 and 11) and tumor type (Questions 12-17). Given that positive and negative categories in Questions 8 and 9 were non-exclusive and addressed in two separate questions, the outcomes were compared as follows: 1) positive versus non-positive (Question 8: TRUE versus FALSE); 2) negative versus non-negative (Question 9: TRUE versus FALSE). The pairwise Fisher Exact Test with false-discovery rate correction for multiple comparison was applied.

## Results

### Dataset Characteristics

The systematic searches retrieved 320 publications for Pembrolizumab, 147 for Cetuximab, and 2055 for Cisplatin. The Cohen’s kappa over all classification tasks was 0.87, indicating a high level of agreement. IRA varied across the questions, ranging from 0.56 to 1.00. Lower agreement with values of <0.70 was observed for the two questions on positive and negative outcomes (0.68 and 0.60), on the tumor type questions for kidney cancer (0.65) and pancreas cancer (0.56) and on two questions regarding therapy sequence (concurrent treatment 0.60; systemic therapy after RT 0.69). A detailed overview of the IRA is provided in **Table 2**. Characteristics of the ground truth datasets are presented in the **Supplementary Table 1**.

**Table 2:**
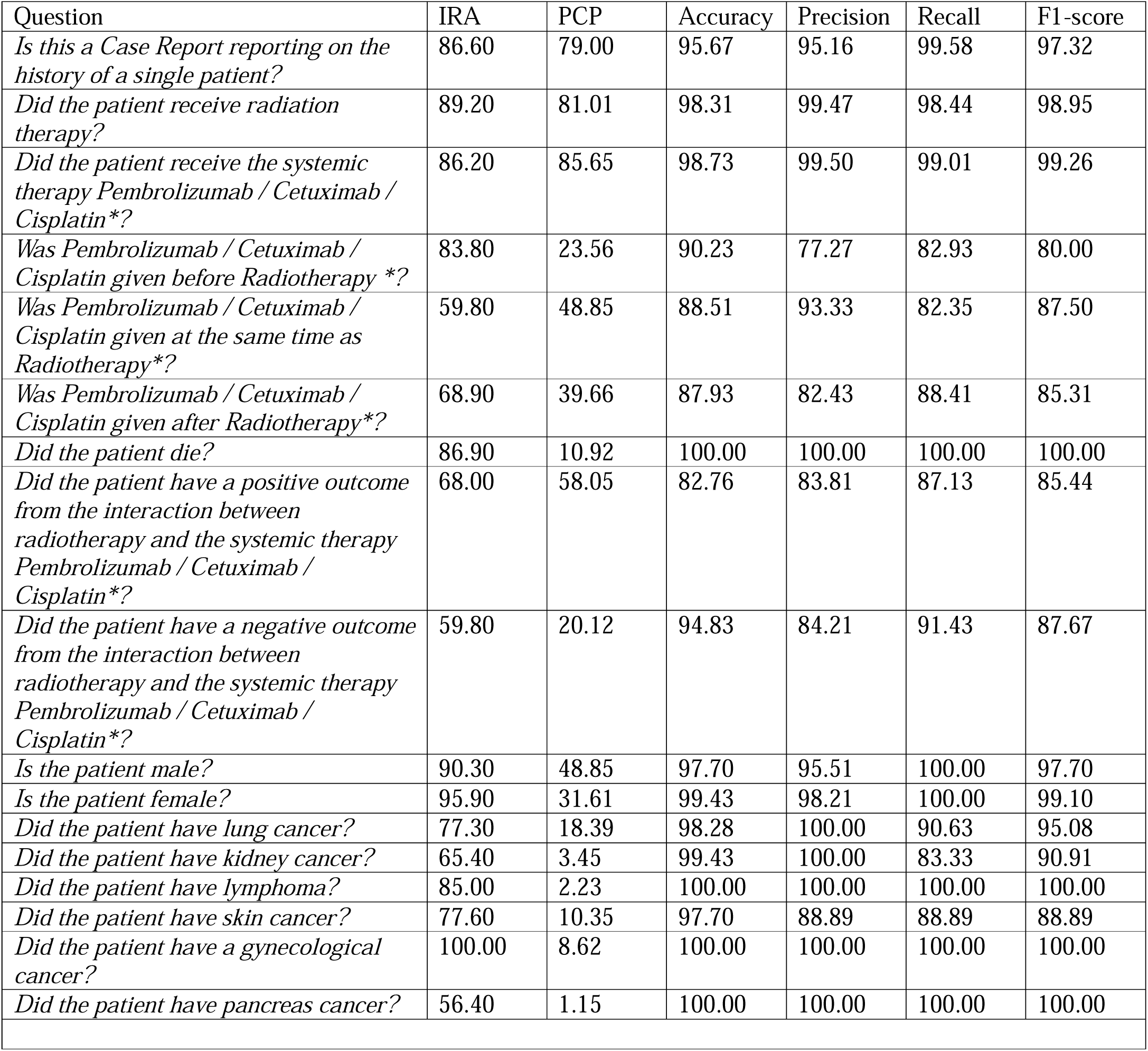
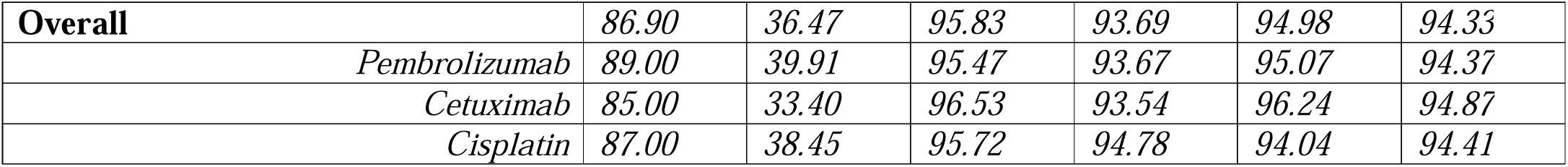
Performance metrics of the LLM-based classification on the seventeen different questions using GPT-OSS-120B. IRA – Inter-rater Agreement calculated using Cohen’s kappa, PCP – Positive Class Prevalence *The question contains only the name of the respective systemic therapy

### LLM Classification Performance on the Ground Truth Datasets

The dataset contained seventeen questions with Boolean classification and had a positive class prevalence of 36.47% (n=1172). The larger model variant GPT-OSS-120B achieved an overall accuracy (=exact match proportion) of 95.83%, precision of 93.69%, recall of 94.98%, and F1-Score of 94.33%. F1-scores>90% were achieved for eleven of the questions.

Lower performance was seen for the three questions regarding treatment sequence (F1-Scores of 80.00%-87.50%) and the questions whether the patient had a positive (F1-Score of 85.44%) or negative (F1-score of 87.67%) outcome (Questions 8 and 9). An overview of the results for GPT-OSS-120B is provided in **Table2**.

IRA significantly correlated with F1-score (ρ=0.48, p=0.028). There were no significant differences in the LLM performance across medications (i.e., Pembrolizumab, Cetuximab and Cisplatin; all p>0.05; **Supplementary Tables 2-4**).

The smaller model variant GPT-OSS-20B achieved similar performance results with an overall accuracy of 95.57%, precision of 91.95%, recall of 96.27% and F1-Score of 94.06% (see **Supplementary Tables 2-4**).

### Aggregate Findings from the Full Datasets

The analysis of the entire datasets (3×100 manually classified publications in ground truth dataset + remaining 2222 publications classified by the LLM) identified 1776 reports as “case reports about the treatment history of a single patient” with 278 reports in the Pembrolizumab dataset, 117 reports in the Cetuximab dataset and 1381 reports in the Cisplatin dataset.

Of these, the patients received both radiation therapy and the respective systemic therapy in 199 (71.58%), 83 (70.94%) and 946 (68.50%) cases.

Across all datasets, case reports where the patient received both RT and the systemic therapy were identified in 56.02 % of all publications. An overview of the proportions is provided in **Figure 2**.

**Figure 2:**
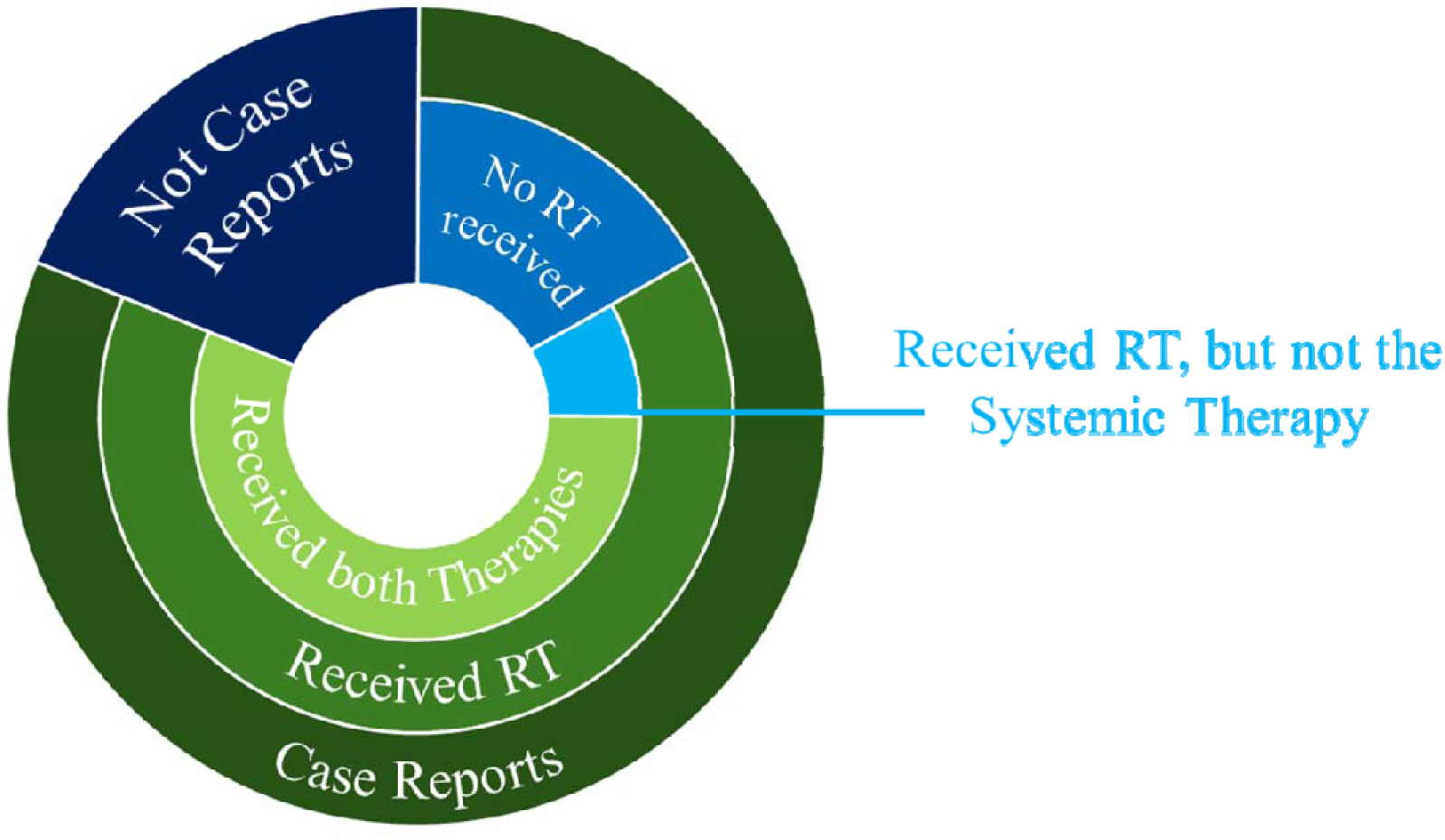
Proportions of publications that were identified as “case reports about the treatment history of a single patient”, as well as of patients that received RT and the systemic therapy across all three datasets. RT – radiation therapy.

The distribution of cancer types included in the analysis showed most of the cases were lung cancer and skin cancer for Pembrolizumab (58 patients and 29 patients) and Cetuximab (4 patients and 7 patients) and lung cancer and gynecological cancer for Cisplatin (127 patients and 102 patients).

Regarding time sequencing of the therapies, for Pembrolizumab most patients received the systemic therapy after completion of RT (120 patients), while for Cetuximab and Cisplatin the therapies were mostly given concurrently (41 patients and 456 patients).

Among patients receiving RT and the systemic therapy, the death of the patient was reported in 24 patients (12.06%) for Pembrolizumab, in 8 patients (9.64%) for Cetuximab and 184 patients (19.45%) for Cisplatin.

For all three systemic therapies, most patients were male (58.64% for Pembrolizumab, 70.91% for Cetuximab, and 59.57% for Cisplatin).

A positive outcome resulting from the interaction of RT and systemic therapy was identified the case reports of 115 patients (70.99%) for Pembrolizumab, of 36 patients (64.54%) for Cetuximab and of 546 patients (64.54%) for Cisplatin.

Negative outcomes resulting from the interaction were identified in the case reports of 46 patients (28.40%) for Pembrolizumab, of 20 patients (34.87%) for Cetuximab, of 295 patients (34.87%) for Cisplatin.

The therapy sequence was significantly associated with the outcome in Cisplatin with more positive outcomes when RT was given concurrently compared to before or after the systemic therapy (each p<0.001). There were also significantly more negative outcomes when RT was given concurrently compared to systemic therapy given before RT (p=0.043). As illustrated in **Figure 3**, this was the only systemic therapy with significant pairwise differences between timing groups.

**Figure 3:**
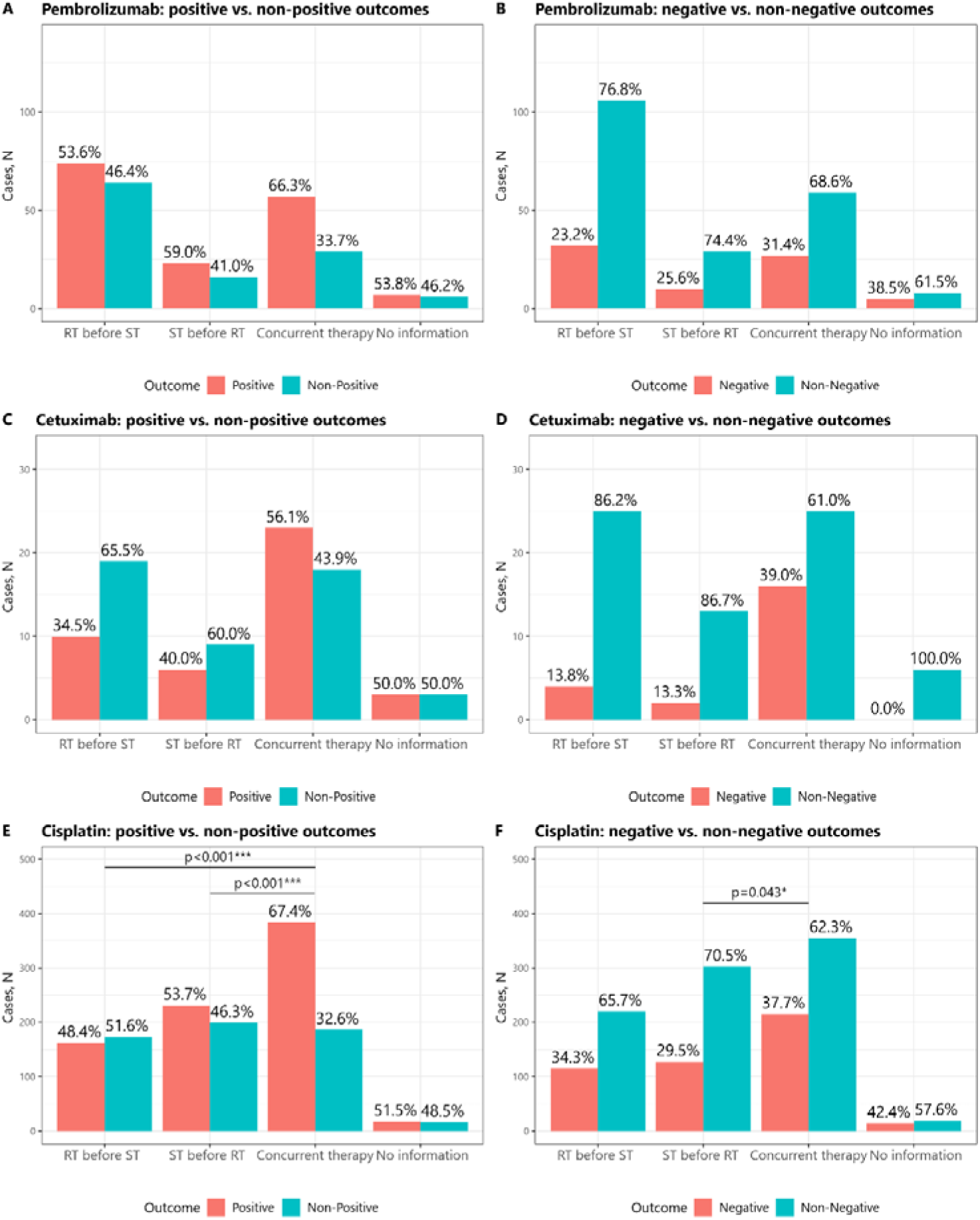
Proportions of outcomes based on the timing of therapy (i.e., RT before ST, ST before RT, concurrent therapy, and no information). A. Pembrolizumab: assessment for positive outcome. B. Pembrolizumab: assessment for negative outcome. C. Cetuximab: assessment for positive outcome. D. Cetuximab: assessment for negative outcome. E. Cisplatin: assessment for positive outcome. F. Cisplatin: assessment for negative outcome. The timing modalities were compared pairwise using McNemar Test with False Discovery Rate correction for multiple comparisons. *p<0.05, **p<0.001, ***p<0.001. RT – radiation therapy, ST – systemic therapy.

Analysis of the data by patient sex (Male, Female, or Unknown) (**Supplementary Figure 1**) or cancer type (**Supplementary Figure 2**), showed no significant association with the outcome.

## Discussion

### LLM-based Analysis of Biomedical Literature, including Case Reports

Our findings demonstrate that LLM-based classification systems can achieve high performance for core clinical tasks in automated curation of scientific case reports involving RT and systemic therapy interactions. This supports their potential utility for high-throughput hypothesis generation and knowledge base construction in oncology (10).

To our knowledge, this is the first study to use LLMs for investigating case reports on combination treatments and their possible interactions. Zhang et al. recently created CaseReportBench (24), which is the first benchmark for information extraction from clinical case reports, containing 138 expert-annotated case reports. Unlike our clearly defined, Boolean structure for data extraction, they used a dense information extraction framework and a more complex multi-domain evaluation approach. There is an increasing number of studies showing high-level performances in information extraction and knowledge synthesis tasks when applying generative AI systems to biomedical literature (25). Case reports remain an underutilized, but highly valuable resource for rare and unique clinical situations. LLMs could be used to better analyzed rare, but important findings from case reports, such as positive or negative outcomes resulting from interactions between RT and systemic therapies.

As we have seen in our study with the Open-Source GPT models, also the smaller 20B version, performed very well with an F1-Score of 94.06%. This is relevant as this model has a size of <15GB and could also be executed on powerful consumer hardware within a local network, which facilitates application to sensitive data (such as clinical cases treated within a healthcare facility).

In our investigation we saw that for Cisplatin, concurrent administration of the systemic therapy with RT (**Figure 3**), is associated with more positive outcomes, which also corresponds to evidence from large clinical trials and recommendations from clinical practice guidelines (26). While this observation is very basic, the fact that this finding aligns with established clinical knowledge serves as a proof-of-concept for the methodology. This alignment functions as a positive control, indicating that the system is capable of capturing real-world clinical patterns. Numerical differences of positive and negative outcomes depending on therapy timing were also observed in Pembrolizumab and Cetuximab, but did not reach statistical significance. Reason for that is most likely the much smaller number of case reports (2055 for Cisplatin compared to 320 for Pembrolizumab and 147 for Cetuximab).

Inferring biological mechanisms from case reports is of course inherently limited. As they describe unique clinical situations without a control group, it is in principle not possible to differentiate the effects of a treatment from confounding variables. Nevertheless, specifically for the use-case of treatment combinations, automated analyses of case reports could help generate new hypotheses on the mechanisms and positive as well as negative interactions.

### Broader Applications and Future Directions

The LLM-based approach for case report analyses could be extended to other questions and domains. The general framework with modular application enables rapid adaptation to new therapies or endpoints. It should be noted that no optimization like training, prompt adaptation or similar was applied in our study. The ground truth dataset was only used to evaluate the performance, not to develop or optimize the system. While the evaluation is crucial to verify the system is working well, the approach itself is general and scalable. It could be directly used in other situations (e.g., other therapy combinations) or with other questions (e.g., regarding specific positive or negative outcomes).

A performant automated classification system applicable to various scenarios would be highly valuable for better understanding and categorization of case reports. It could be used to create a detailed database that is automatically updated without the need for human curation. Such a database would enable clinicians to easily find cases relevant to their own patients presenting with a rare clinical scenario. Clinicians and researchers could benefit much more from the experiences of previously treated patients with rare disease situations.

Future efforts will include application to additional systemic therapies, extension to more detailed clinical data collection, and if needed possibly optimization of performance (e.g., using prompt optimization or model fine-tuning). Furthermore, future studies on case reports may include analyses of the full text documents, potentially using Vision Language Models (VLM) for extraction of information not only from free text but from images and graphics (such as Figures or photos).

### Limitations

Despite promising results, several challenges remain. The LLMs performed slightly worse on questions related to time sequencing of RT and systemic therapy as well as on the highly relevant questions of positive and negative outcomes resulting from therapy interactions. We hypothesize that the reason for this lies in the increased ambiguity of these questions. As we have also seen in the IRA, there was a lower level of agreement between the two human researchers on these questions. Since we did not provide a detailed definition of what is considered a “positive” or “negative” outcome, this is not surprising. Clear definitions of questions are crucial to reduce ambiguities and obtain clear answers (both for human-based as well as LLM-based classification tasks).

Another limitation of our study is its descriptive scope, which is a consequence of our methodological design. We used a limited set of 17 well-defined Boolean questions. Asking whether the patient had “Lung Cancer (TRUE/FALSE)”, “Skin Cancer (TRUE/FALSE)”, “Kidney Cancer (TRUE/FALSE)” etc. instead of just asking “What cancer did the patient have? (Open question)” might appear time-consuming and unnecessary. Yet, there are many special clinical scenarios (particularly in case reports) and open-ended questions (or questions with multiple categories), that might be difficult to answer (e.g., patient with several cancer diseases).

While this pilot study does not provide new deep insights into interactions between RT and systemic therapies, future work will expand the data extraction schema to include more detailed questions, such as treatment parameters, specific toxicity profiles, and detailed patient outcomes. Using such a strategy, LLMs may help to obtain a better overview and understanding of case reports about interactions of different oncological therapies.

## Conclusion

LLM-based systems for classifying scientific case reports on specific questions, such as interactions between RT and systemic therapies, have huge potential. Despite some limitations in questions requiring more nuanced interpretation, the high performance and scalability of this approach suggest high value in automating knowledge extraction from case reports. This capability can be used in knowledge synthesis and hypothesis generation to assist in the development of clinical guidelines for complex treatment scenarios, ultimately enhancing the utility of clinical case reports.

## List of abbreviations

AI: Artificial Intelligence
GB: Gigabyte
GPT: Generative Pretrained Transformer
IRA: Inter-rater Agreement
JSON: JavaScript Object Notation
LLM: Large Language Model
MeSH: Medical Subject Headings
NLP: Natural Language Processing
PD-L1: Programmed Death Ligand 1
RT: Radiotherapy
ST: Systemic Therapy
VLM: Vision Language Model

## Acknowledgement

Not applicable.

## Statement of Ethics

Not applicable.

## Conflict of Interest Statement

Dr. Cihoric is a technical lead for the *SmartOncology* project and medical advisor for Wemedoo AG, Steinhausen AG, Switzerland. The authors declare no other conflicts of interest.

## Funding Sources

No funding was received for this work.

## Author contributions

Conceptualization – FD, NC, MS

Data Collection –TB, AH, FD

Writing, original draft preparation – FD, NC

Writing, illustrations – FD, IF

Statistical analysis – FD, IF

Writing, review and editing – All authors

## Data availability statement

The source code for the project as well as the .JSON files containing the relevant data about the data structure with classification topics, categories and prompts are provided at GitHub (21).

