## Supplementary File 1 for "LLM-Based Classification of Case Report Abstracts: A Pilot Study on Interactions between Radiotherapy and Systemic Therapies"

The following contains the details on creation of the systematic queries to obtain relevant case reports from PubMed.

**Radiotherapy**

**MeSH Terms:**

- Radiotherapies
- Radiation Therapy
- Radiation Therapies
- Therapies, Radiation
- Therapy, Radiation
- Radiation Treatment
- Radiation Treatments
- Treatment, Radiation
- Radiotherapy, Targeted
- Radiotherapies, Targeted
- Targeted Radiotherapies
- Targeted Radiation Therapy
- Radiation Therapies, Targeted
- Targeted Radiation Therapies
- Therapies, Targeted Radiation
- Therapy, Targeted Radiation
- Radiation Therapy, Targeted
- Targeted Radiotherapy

(https://www.ncbi.nlm.nih.gov/mesh/68011878)

**Pembrolizumab**

**MeSH Terms:**

- MK-3475
- Keytruda
- lambrolizumab
- SCH-900475

(https://www.ncbi.nlm.nih.gov/mesh/?term=Pembrolizumab)

**Query:**

((("Radiotherapies" OR "Radiation Therapy" OR "Radiation Therapies" OR "Therapies, Radiation" OR "Therapy, Radiation" OR "Radiation Treatment" OR "Radiation Treatments" OR "Treatment, Radiation" OR "Radiotherapy, Targeted" OR "Radiotherapies, Targeted" OR "Targeted Radiotherapies" OR "Targeted Radiotherapy" OR "Targeted Radiation Therapy" OR "Radiation Therapies, Targeted" OR "Targeted Radiation Therapies" OR "Therapies, Targeted Radiation" OR "Therapy, Targeted Radiation" OR "Radiation Therapy, Targeted") AND ("Pembrolizumab" OR "SCH-900475" OR "lambrolizumab" OR "MK-3475" OR "Keytruda") )) AND ("1800/01/01"[Date - Publication] : "2024/12/31"[Date - Publication])

**Link:** <https://pubmed.ncbi.nlm.nih.gov/?term=%28%28%28%22Radiotherapies%22+OR+%22Radiation+Therapy%22+OR+%22Radiation+Therapies%22+OR+%22Therapies%2C+Radiation%22+OR+%22Therapy%2C+Radiation%22+OR+%22Radiation+Treatment%22+OR+%22Radiation+Treatments%22+OR+%22Treatment%2C+Radiation%22+OR+%22Radiotherapy%2C+Targeted%22+OR+%22Radiotherapies%2C+Targeted%22+OR+%22Targeted+Radiotherapies%22+OR+%22Targeted+Radiotherapy%22+OR+%22Targeted+Radiation+Therapy%22+OR+%22Radiation+Therapies%2C+Targeted%22+OR+%22Targeted+Radiation+Therapies%22+OR+%22Therapies%2C+Targeted+Radiation%22+OR+%22Therapy%2C+Targeted+Radiation%22+OR+%22Radiation+Therapy%2C+Targeted%22%29+AND+%28%22Pembrolizumab%22+OR+%22SCH-900475%22+OR+%22lambrolizumab%22+OR+%22MK-3475%22+OR+%22Keytruda%22%29+%29%29+AND+%28%221800%2F01%2F01%22%5BDate+-+Publication%5D+%3A+%222024%2F12%2F31%22%5BDate+-+Publication%5D%29&filter=pubt.casereports>

**Cetuximab**

**MeSH Terms:**

- IMC C225
- MAb C225
- C225
- IMC-C225
- Erbitux

(https://www.ncbi.nlm.nih.gov/mesh/2009883)

**Query:**

((("Radiotherapies" OR "Radiation Therapy" OR "Radiation Therapies" OR "Therapies, Radiation" OR "Therapy, Radiation" OR "Radiation Treatment" OR "Radiation Treatments" OR "Treatment, Radiation" OR "Radiotherapy, Targeted" OR "Radiotherapies, Targeted" OR "Targeted Radiotherapies" OR "Targeted Radiotherapy" OR "Targeted Radiation Therapy" OR "Radiation Therapies, Targeted" OR "Targeted Radiation Therapies" OR "Therapies, Targeted Radiation" OR "Therapy, Targeted Radiation" OR "Radiation Therapy, Targeted") AND ("Cetuximab" OR "Erbitux" OR "IMC C225" OR "IMC-C225" OR "MAb C225" OR "C225") )) AND ("1800/01/01"[Date - Publication] : "2024/12/31"[Date - Publication])

**Link:** <https://pubmed.ncbi.nlm.nih.gov/?term=%28%28%28%22Radiotherapies%22+OR+%22Radiation+Therapy%22+OR+%22Radiation+Therapies%22+OR+%22Therapies%2C+Radiation%22+OR+%22Therapy%2C+Radiation%22+OR+%22Radiation+Treatment%22+OR+%22Radiation+Treatments%22+OR+%22Treatment%2C+Radiation%22+OR+%22Radiotherapy%2C+Targeted%22+OR+%22Radiotherapies%2C+Targeted%22+OR+%22Targeted+Radiotherapies%22+OR+%22Targeted+Radiotherapy%22+OR+%22Targeted+Radiation+Therapy%22+OR+%22Radiation+Therapies%2C+Targeted%22+OR+%22Targeted+Radiation+Therapies%22+OR+%22Therapies%2C+Targeted+Radiation%22+OR+%22Therapy%2C+Targeted+Radiation%22+OR+%22Radiation+Therapy%2C+Targeted%22%29+AND+%28%22Cetuximab%22+OR+%22Erbitux%22+OR+%22IMC+C225%22+OR+%22IMC-C225%22+OR+%22MAb+C225%22+OR+%22C225%22%29+%29%29+AND+%28%221800%2F01%2F01%22%5BDate+-+Publication%5D+%3A+%222024%2F12%2F31%22%5BDate+-+Publication%5D%29&filter=pubt.casereports>

**Cisplatin**

**MeSH Terms:**

- cis-Diamminedichloroplatinum
- cis Diamminedichloroplatinum
- cis-Diamminedichloroplatinum(II)
- cis-Dichlorodiammineplatinum(II)
- cis-Platinum
- cis Platinum
- Dichlorodiammineplatinum
- Platinum Diamminodichloride
- Diamminodichloride, Platinum
- Biocisplatinum
- Platidiam
- Platino
- Platinol
- NSC-119875

(https://www.ncbi.nlm.nih.gov/mesh/68002945)

**Query:**

((("Radiotherapies" OR "Radiation Therapy" OR "Radiation Therapies" OR "Therapies, Radiation" OR "Therapy, Radiation" OR "Radiation Treatment" OR "Radiation Treatments" OR "Treatment, Radiation" OR "Radiotherapy, Targeted" OR "Radiotherapies, Targeted" OR "Targeted Radiotherapies" OR "Targeted Radiotherapy" OR "Targeted Radiation Therapy" OR "Radiation Therapies, Targeted" OR "Targeted Radiation Therapies" OR "Therapies, Targeted Radiation" OR "Therapy, Targeted Radiation" OR "Radiation Therapy, Targeted") AND ("cisplatin" OR "cis-Diamminedichloroplatinum(II)" OR "Platinum Diamminodichloride" OR "Diamminodichloride, Platinum" OR "cis-Platinum" OR "cis Platinum" OR "Dichlorodiammineplatinum" OR "cis-Diamminedichloroplatinum" OR "cis Diamminedichloroplatinum" OR "cis-Dichlorodiammineplatinum(II)" OR "NSC-119875" OR "Platino" OR "Platinol" OR "Biocisplatinum" OR "Platidiam") )) AND ("1800/01/01"[Date - Publication] : "2024/12/31"[Date - Publication])

**Link:**

<https://pubmed.ncbi.nlm.nih.gov/?term=%28%28%28%22Radiotherapies%22+OR+%22Radiation+Therapy%22+OR+%22Radiation+Therapies%22+OR+%22Therapies%2C+Radiation%22+OR+%22Therapy%2C+Radiation%22+OR+%22Radiation+Treatment%22+OR+%22Radiation+Treatments%22+OR+%22Treatment%2C+Radiation%22+OR+%22Radiotherapy%2C+Targeted%22+OR+%22Radiotherapies%2C+Targeted%22+OR+%22Targeted+Radiotherapies%22+OR+%22Targeted+Radiotherapy%22+OR+%22Targeted+Radiation+Therapy%22+OR+%22Radiation+Therapies%2C+Targeted%22+OR+%22Targeted+Radiation+Therapies%22+OR+%22Therapies%2C+Targeted+Radiation%22+OR+%22Therapy%2C+Targeted+Radiation%22+OR+%22Radiation+Therapy%2C+Targeted%22%29+AND+%28%22cisplatin%22+OR+%22cis-Diamminedichloroplatinum%28II%29%22+OR+%22Platinum+Diamminodichloride%22+OR+%22Diamminodichloride%2C+Platinum%22+OR+%22cis-Platinum%22+OR+%22cis+Platinum%22+OR+%22Dichlorodiammineplatinum%22+OR+%22cis-Diamminedichloroplatinum%22+OR+%22cis+Diamminedichloroplatinum%22+OR+%22cis-Dichlorodiammineplatinum%28II%29%22+OR+%22NSC-119875%22+OR+%22Platino%22+OR+%22Platinol%22+OR+%22Biocisplatinum%22+OR+%22Platidiam%22%29+%29%29+AND+%28%221800%2F01%2F01%22%5BDate+-+Publication%5D+%3A+%222024%2F12%2F31%22%5BDate+-+Publication%5D%29&filter=pubt.casereports>
