## Supplementary File 2 for "LLM-Based Classification of Case Report Abstracts: A Pilot Study on Interactions between Radiotherapy and Systemic Therapies"

### TRIPOD+LLM Checklist

| Section / Topic | Item Number | Checklist Item | Research Design | LLM Task | Reported on Page |
|---|---|---|---|---|---|
| **Abstract** | | | | | |
| **Title** | 2a | Identify the study as developing, fine-tuning, and/or evaluating the performance of an LLM, specifying the task, the target population, and the outcome to be predicted. | All | All | 1 |
| **Abstract** | 2b | Provide a brief explanation of the healthcare context, use case and rationale for developing or evaluating the performance of an LLM. | E,H | All | 1 |
| **Objectives** | 2c | Specify the study objectives, including whether the study describes LLMs development, tuning, and/or evaluation | All | All | 1 |
| **Methods** | 2d | Describe the key elements of the study setting. | All | All | 1 |
| | 2e | Detail all data used in the study, specify data splits and any selective use of data. | M,D,E | All | 1 |
| | 2f | Specify the name and version of LLM used. | All | All | 1 |
| | 2g | Briefly summarize the LLM-building steps, including any fine-tuning, reward modeling, reinforcement learning with human feedback (RLHF), etc. | M,D | All | Not Required |
| | 2h | Describe the specific tasks performed by the LLMs (e.g., medical QA, summarization, extraction), highlighting key inputs and outputs used in the final LLM. | All | All | 1 |
| | 2i | Specify the evaluation datasets/populations used, including the endpoint evaluated, and detail whether this information was held out during training/tuning where relevant, and what measure(s) were used to evaluate LLM performance. | All | All | 1 |
| **Results** | 2j | Give an overall report and interpretation of the main results. | All | All | 1 |
| **Discussion** | 2k | Explicitly state any broader implications or concerns that have arisen in light of these results. | All | All | 1 |
| **Other** | 2l | Give the registration number and name of the registry or repository (if relevant). | H | All | Not Required |
| **Introduction** | | | | | |

| Section / Topic | Item Number | Checklist Item | Research Design | LLM Task | Reported on Page |
|---|---|---|---|---|---|
| **Background** | 3a | Explain the healthcare context / use case (e.g., administrative, diagnostic, therapeutic, clinical workflow) and rationale for developing or evaluating the LLM, including references to existing approaches and models. | All | All | 2, 3 |
| | 3b | Describe the target population and the intended use of the LLM in the context of the care pathway, including its intended users in current gold standard practices (e.g., healthcare professionals, patients, public, or administrators). | E,H | All | 2, 3 |
| **Objectives** | 4 | Specify the study objectives, including whether the study describes the initial development, fine-tuning, or validation of an LLM (or multiple stages). | All | All | 3 |
| **Methods** | | | | | |
| **Data** | 5a | Describe the sources of data separately for the training, tuning, and/or evaluation datasets and the rationale for using these data (e.g., web corpora, clinical research/trial data, EHR data). | All | All | 3, 4, 5 |
| | 5b | Describe the relevant data points and provide a quantitative and qualitative description of their distribution and other relevant descriptors of the dataset (e.g., source, languages, countries of origin) | All | All | 3 |
| | 5c | Specifically state the date of the oldest and newest item of text used in the development process (training, fine-tuning, reward modeling) and in the evaluation datasets. | M,D,E,H | All | N/A |
| | 5d | Describe any data pre-processing and quality checking, including whether this was similar across text corpora, institutions, and relevant sociodemographic groups. | All | All | 3, 4, 5 |
| | 5e | Describe how missing and imbalanced data were handled and provide reasons for omitting any data. | M,D,E | All | N/A |

| Section / Topic | Item Number | Checklist Item | Research Design | LLM Task | Reported on Page |
|---|---|---|---|---|---|
| Analytical Methods | 6a | Report the LLM name, version, and last date of training or use during inference. | All | All | 5 |
| | 6b | Specify the type of LLM architecture, and LLM building steps, including any hyperparameter tuning (e.g., temperature, length limits, penalties), prompt engineering, and any inference settings (e.g., seed, temperature, max token length) as relevant. | M,D,E | All | 6, GitHub |
| | 6c | Report details of LLM development process from text input to outcome generation, such as training, fine-tuning procedures, and alignment strategy (e.g., reinforcement learning, direct preference optimization, etc.) and alignment goals (e.g., helpfulness, honesty, harmlessness, etc.). | M,D | All | Not Required |
| | 6d | Specify the initial and post-processed output of the LLM (e.g., probabilities, classification, unstructured text). | All | All | 6, GitHub |
| | 6e | Provide details and rationale for any classification and how the probabilities were determined and thresholds identified. | All | C,OF | Not Required |
| | 6f | Include metrics that capture the quality of generative outputs, such as consistency, relevance, and accuracy, compared to gold standards. | All | QA,IR,DG,SS,MT | Not Required |
| | 6g | Report the outcome metrics' relevance to downstream task at deployment time and correlation of metric to human evaluation of the text for the intended use. | E,H | All | N/A |
| LLM Output | 7a | Clearly define the outcome, how the LLM predictions were calculated (e.g., formula, code, object, API), and evaluation metrics. | E,H | All | 6, GitHub |
| | 7b | If outcome assessment requires subjective interpretation, describe the qualifications of the assessors, any instructions provided, relevant information on demographics of the assessors, and inter-assessor agreement. | All | All | N/A |
| | 7c | Specify how performance was compared to other LLMs, humans, and other benchmarks or standards. | All | All | 6 |

| Section / Topic | Item Number | Checklist Item | Research Design | LLM Task | Reported on Page |
|---|---|---|---|---|---|
| **Annotation** | 8a | If annotation was done, report how text was labeled, including providing specific annotation guidelines with examples. | All | All | 5 |
| | 8b | If annotation was done, report how many annotators labeled the dataset(s), including the proportion of data in each dataset that were annotated by more than 1 annotator. | All | All | 5 |
| | 8c | If annotation was done, provide information on the background and experience of the annotators, and the inter-annotator agreement. | All | All | 5 |
| **Prompting** | 9a | If research involved prompting LLMs, provide details on the processes used during prompt design, curation, and selection. | All | All | 5 |
| | 9b | If research involved prompting LLMs, report what data were used to develop the prompts. | All | All | 5 |
| **Summarization** | 10 | Describe any preprocessing of the data before summarization. | All | SS | Not Required |
| **Instruction Tuning / Alignment** | 11 | If instruction tuning/alignment strategies were used, what were the instructions and interface used for evaluation, and what were the characteristics of the populations doing evaluation? | M,D | All | Not Required |
| **Compute** | 12 | Report compute, or proxies thereof (e.g., time on what and how many machines, cost on what and how many machines, inference time, floating-point operations per second (FLOPs)), required to carry out methods. | M,D,E | All | 6 |
| **Ethics Approval** | 13 | Name the institutional research board or ethics committee that approved the study and describe the participant-informed consent or the ethics committee waiver of informed consent. | All | All | N/A |

| Section / Topic | Item Number | Checklist Item | Research Design | LLM Task | Reported on Page |
|---|---|---|---|---|---|
| **Open Science** | 14a | Give the source of funding and the role of the funders for the present study. | All | All | N/A |
| | 14b | Declare any conflicts of interest and financial disclosures for all authors. | All | All | 12 |
| | 14c | Indicate where the study protocol can be accessed or state that a protocol was not prepared. | H | All | Not Required |
| | 14d | Provide registration information for the study, including register name and registration number, or state that the study was not registered. | H | All | Not Required |
| | 14e | Provide details of the availability of the study data. | All | All | 6 |
| | 14f | Provide details of the availability of the code to reproduce the study results. | All | All | 6 |
| **Public Involvement** | 15 | Provide details of any patient and public involvement during the design, conduct, reporting, interpretation, or dissemination of the study or state no involvement. | H | All | Not Required |
| **Results** | | | | | |
| **Participants** | 16a | When using patient/EHR data, describe the flow of text/EHR/patient data through the study, including the number of documents/questions/participants with and without the outcome/label and follow-up time. | E,H | All | N/A |
| | 16b | When using patient/EHR data, report the characteristics overall and, for each data source or setting, and for development/evaluation splits, including the key dates, key predictors, and sample size. | E,H | All | N/A |
| | 16c | For LLM evaluation, show a comparison of the distribution of important predictors between development and evaluation data. | E,H | All | N/A |
| | 16d | When using patient/EHR data, specify the number of participants and outcome events in each analysis (e.g., for LLM development, hyperparameter tuning, LLM evaluation). | E,H | All | N/A |
| **Performance** | 17 | Report LLM performance according to pre-specified metrics (see item 7a) and/or human evaluation (see item 7d). | All | All | 6, 7, 8 |
| **LLM Updating** | 18 | If applicable, report the results from any LLM updating, including the updated LLM and subsequent performance. | All | All | N/A |
| **Discussion** | | | | | |

| Section / Topic | Item Number | Checklist Item | Research Design | LLM Task | Reported on Page |
|---|---|---|---|---|---|
| **Interpretation** | 19a | Give an overall interpretation of the main results, including issues of fairness in the context of the objectives and previous studies. | All | All | 9, 10 |
| **Limitations** | 19b | Discuss any limitations of the study and their effects on any biases, statistical uncertainty, and generalizability. | All | All | 11 |
| **Usability of the LLM in context** | 19c | Describe any known challenges in using data for the specified task and domain context with reference to representation, missingness, harmonization, and bias. | E,H | All | 11 |
| | 19d | Define the intended use for the implementation under evaluation, including the intended input, end-user, level of autonomy/human oversight. | E,H | All | 10, 11 |
| | 19e | If applicable, describe how poor quality or unavailable input data should be assessed and handled when implementing the LLM, i.e., what is the usability of the LLM in the context of current clinical care. | E,H | All | N/A |
| | 19f | If applicable, specify whether users will be required to interact in the handling of the input data or use of the LLM, and what level of expertise is required of users. | E,H | All | N/A |
| | 19g | Discuss any next steps for future research, with a specific view to applicability and generalizability of the LLM. | All | All | 11 |
