## Supplementary Figures for "LLM-Based Classification of Case Report Abstracts: A Pilot Study on Interactions between Radiotherapy and Systemic Therapies"

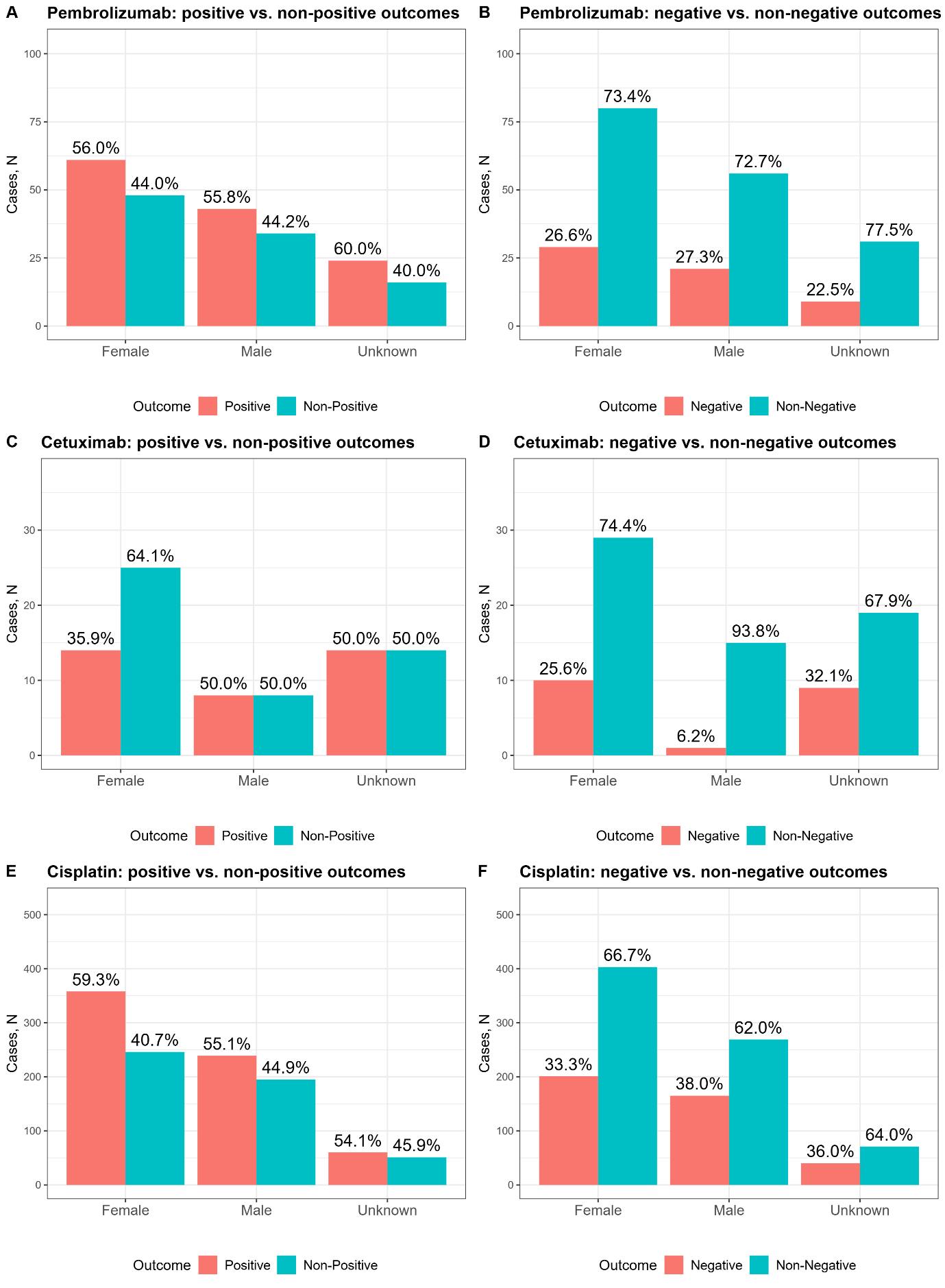

Supplementary Figure 1: Proportions of outcomes based on patient sex. A. Pembrolizumab: assessment for positive outcome. B. Pembrolizumab: assessment for negative outcome . C. Cetuximab: assessment for positive outcome . D. Cetuximab: assessment for negative outcome . E. Cisplatin: assessment for positive outcome . F. Cisplatin: assessment for negative outcome. The patient sex categories were compared pairwise using McNemar Test with False Discovery Rate correction for multiple comparisons. *p<0.05, **p<0.001, ***p<0.001.

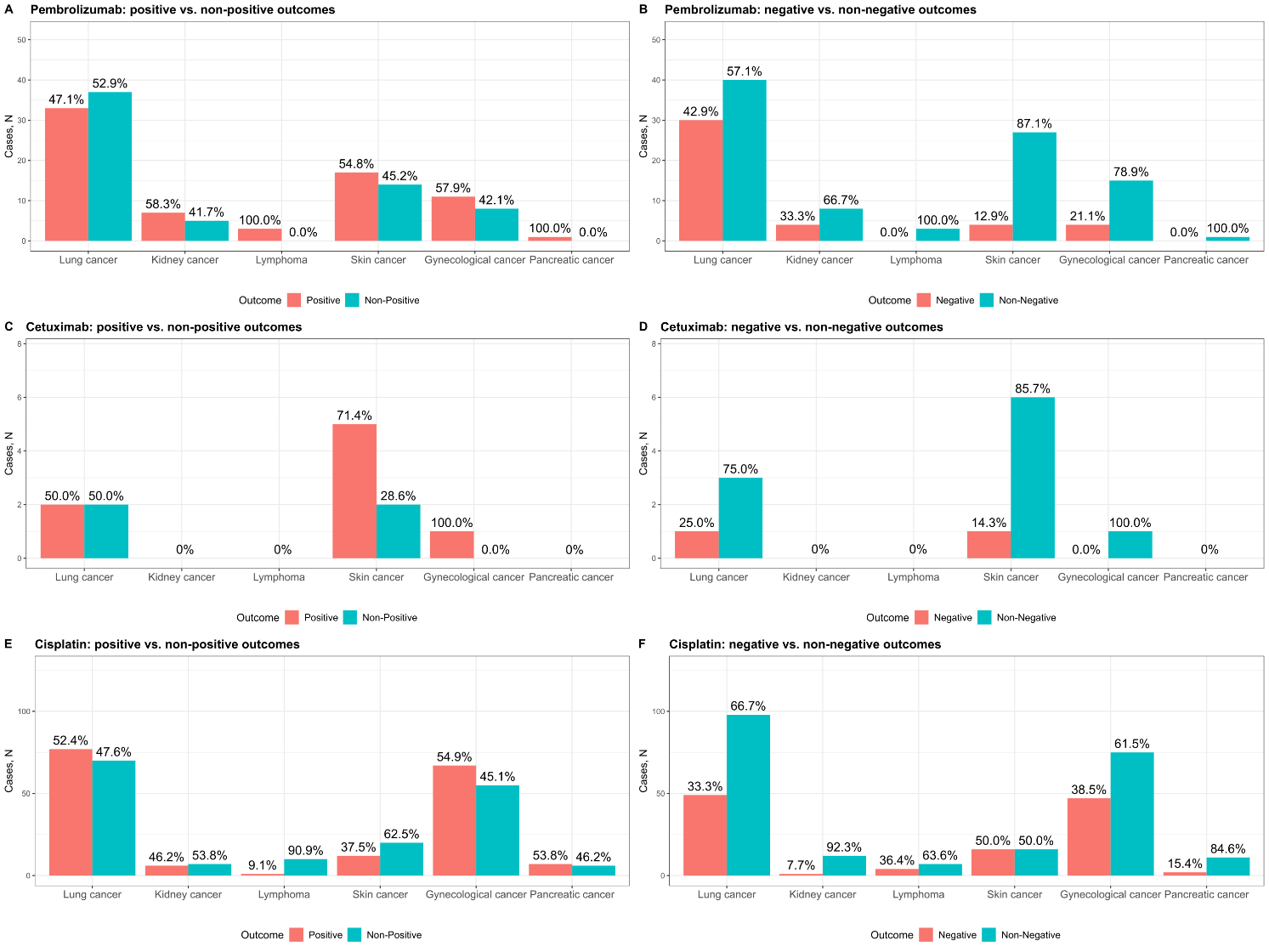

Supplementary Figure 2: Proportions of outcomes based on patient cancer type. A. Pembrolizumab: assessment for positive outcome. B. Pembrolizumab: assessment for negative outcome . C. Cetuximab: assessment for positive outcome . D. Cetuximab: assessment for negative outcome . E. Cisplatin: assessment for positive outcome . F. Cisplatin: assessment for negative outcome. The cancer type categories were compared pairwise using McNemar Test with False Discovery Rate correction for multiple comparisons. *p<0.05, **p<0.001, ***p<0.001.
