## Supplementary Tables for "LLM-Based Classification of Case Report Abstracts: A Pilot Study on Interactions between Radiotherapy and Systemic Therapies"

| **Question** | | | **Pembrolizumab** | **Cetuximab** | **Cisplatin** |
| --- | --- | --- | --- | --- | --- |
| *Case Report reporting on the history of a single patient* | | | 87 | 76 | 74 |
|  | *Patient received RT* | | 70 | 57 | 65 |
|  | *Patient received the systemic therapy* | | 75 | 71 | 57 |
|  | *Patient received both therapies* | | 64 | 56 | 54 |
|  |  | *Systemic therapy was given before RT* | 16 | 8 | 17 |
|  |  | *Systemic therapy was given together with RT* | 24 | 28 | 33 |
|  |  | *Systemic therapy was given after RT* | 36 | 18 | 15 |
|  |  | *Patient died* | 6 | 5 | 8 |
|  |  | *Patient had a positive outcome from the interaction between RT and the systemic therapy* | 41 | 25 | 35 |
|  |  | *Patient had a negative outcome from the interaction between RT and the systemic therapy* | 8 | 15 | 12 |
|  |  | *Patient is male* | 35 | 25 | 25 |
|  |  | *Patient is female* | 22 | 10 | 23 |
|  |  | *Patient has lung cancer* | 19 | 3 | 10 |
|  |  | *Patient has kidney cancer* | 4 | 0 | 2 |
|  |  | *Patient has lymphoma* | 3 | 0 | 1 |
|  |  | *Patient has skin cancer* | 12 | 4 | 2 |
|  |  | *Patient has gynecological cancer* | 8 | 1 | 6 |
|  |  | *Patient had pancreas cancer* | 1 | 0 | 1 |

Supplementary Table 1: Characteristics of the ground truth datasets based on the questions that were answered with “TRUE”.

| **Question** | **Accuracy** | | **Precision** | | **Recall** | | **F1-score** | |
| --- | --- | --- | --- | --- | --- | --- | --- | --- |
|  | GPT-OSS 20B | GPT-OSS 120B | GPT-OSS 20B | GPT-OSS 120B | GPT-OSS 20B | GPT-OSS 120B | GPT-OSS 20B | GPT-OSS 120B |
| *Is this a Case Report reporting on the history of a single patient?* | 96.0% | 96.0% | 95.6% | 95.6% | 100.0% | 100.0% | 97.8% | 97.8% |
| *Did the patient receive radiation therapy?* | 100.0% | 100.0% | 100.0% | 100.0% | 100.0% | 100.0% | 100.0% | 100.0% |
| *Did the patient receive the systemic therapy Pembrolizumab?* | 97.7% | 100.0% | 97.4% | 100.0% | 100.0% | 100.0% | 98.7% | 100.0% |
| *Was Pembrolizumab given before Radiotherapy?* | 87.5% | 89.1% | 70.0% | 84.6% | 87.5% | 68.8% | 77.8% | 75.9% |
| *Was Pembrolizumab given at the same time as Radiotherapy?* | 93.8% | 85.9% | 95.5% | 85.7% | 87.5% | 75.0% | 91.3% | 80.0% |
| *Was Pembrolizumab given after RT* ********?* | 82.8% | 79.7% | 80.5% | 76.7% | 91.7% | 91.7% | 85.7% | 83.5% |
| *Did the patient die?* | 96.9% | 98.4% | 83.3% | 100.0% | 83.3% | 83.3% | 83.3% | 90.9% |
| *Did the patient have a positive outcome from the interaction between radiotherapy and the systemic therapy Pembrolizumab?* | 79.7% | 79.7% | 80.4% | 80.4% | 90.2% | 90.2% | 85.1% | 85.1% |
| *Did the patient have a negative outcome from the interaction between radiotherapy and the systemic therapy Pembrolizumab?* | 93.8% | 95.3% | 75.0% | 77.8% | 75.0% | 87.5% | 75.0% | 82.4% |
| *Is the patient male?* | 100.0% | 100.0% | 100.0% | 100.0% | 100.0% | 100.0% | 100.0% | 100.0% |
| *Is the patient female?* | 98.4% | 100.0% | 95.7% | 100.0% | 100.0% | 100.0% | 97.8% | 100.0% |
| *Did the patient have lung cancer?* | 98.4% | 98.4% | 100.0% | 100.0% | 94.7% | 94.7% | 97.3% | 97.3% |
| *Did the patient have kidney cancer?* | 98.4% | 98.4% | 100.0% | 100.0% | 75.0% | 75.0% | 85.7% | 85.7% |
| *Did the patient have lymphoma?* | 100.0% | 100.0% | 100.0% | 100.0% | 100.0% | 100.0% | 100.0% | 100.0% |
| *Did the patient have skin cancer?* | 98.4% | 98.4% | 92.3% | 100.0% | 100.0% | 91.7% | 96.0% | 95.7% |
| *Did the patient have a gynecological cancer?* | 100.0% | 100.0% | 100.0% | 100.0% | 100.0% | 100.0% | 100.0% | 100.0% |
| *Did the patient have pancreas cancer?* | 100.0% | 100.0% | 100.0% | 100.0% | 100.0% | 100.0% | 100.0% | 100.0% |
| **Overall** | **95.6** | **95.5** | **92.8** | **93.7** | **96.3** | **95.1** | **94.5** | **94.4** |

Supplementary Table 2: Performance metrics of the LLM-based classification on the seventeen different questions for the Pembrolizumab dataset.

| **Question** | **Accuracy** | | **Precision** | | **Recall** | | **F1-score** | |
| --- | --- | --- | --- | --- | --- | --- | --- | --- |
|  | GPT-OSS 20B | GPT-OSS 120B | GPT-OSS 20B | GPT-OSS 120B | GPT-OSS 20B | GPT-OSS 120B | GPT-OSS 20B | GPT-OSS 120B |
| *Is this a Case Report reporting on the history of a single patient?* | 94.0% | 94.0% | 92.7% | 92.7% | 100.0% | 100.0% | 96.2% | 96.2% |
| *Did the patient receive radiation therapy?* | 100.0% | 98.7% | 100.0% | 100.0% | 100.0% | 98.2% | 100.0% | 99.1% |
| *Did the patient receive the systemic therapy Cetuximab?* | 97.4% | 97.4% | 98.6% | 98.6% | 98.6% | 98.6% | 98.6% | 98.6% |
| *Was Cetuximab given before Radiotherapy?* | 80.4% | 92.9% | 41.2% | 70.0% | 87.5% | 87.5% | 56.0% | 77.8% |
| *Was Cetuximab given at the same time as Radiotherapy?* | 91.1% | 96.4% | 87.1% | 96.4% | 96.4% | 96.4% | 91.5% | 96.4% |
| *Was Cetuximab given after RT?* | 83.9% | 87.5% | 71.4% | 82.4% | 83.3% | 77.8% | 76.9% | 80.0% |
| *Did the patient die?* | 98.2% | 98.2% | 83.3% | 83.3% | 100.0% | 100.0% | 90.9% | 90.9% |
| *Did the patient have a positive outcome from the interaction between radiotherapy and the systemic therapy Cetuximab?* | 87.5% | 89.3% | 84.6% | 88.0% | 88.0% | 88.0% | 86.3% | 88.0% |
| *Did the patient have a negative outcome from the interaction between radiotherapy and the systemic therapy Cetuximab?* | 96.4% | 100.0% | 88.2% | 100.0% | 100.0% | 100.0% | 93.8% | 100.0% |
| *Is the patient male?* | 94.6% | 94.6% | 89.3% | 89.3% | 100.0% | 100.0% | 94.3% | 94.3% |
| *Is the patient female?* | 98.2% | 98.2% | 90.9% | 90.9% | 100.0% | 100.0% | 95.2% | 95.2% |
| *Did the patient have lung cancer?* | 96.4% | 96.4% | 100.0% | 100.0% | 33.3% | 33.3% | 50.0% | 50.0% |
| *Did the patient have kidney cancer?* | 100.0% | 100.0% | - | - | - | - | - | - |
| *Did the patient have lymphoma?* | 100.0% | 100.0% | - | - | - | - | - | - |
| *Did the patient have skin cancer?* | 98.2% | 98.2% | 80.0% | 80.0% | 100.0% | 100.0% | 88.9% | 88.9% |
| *Did the patient have a gynecological cancer?* | 100.0% | 100.0% | 100.0% | 100.0% | 100.0% | 100.0% | 100.0% | 100.0% |
| *Did the patient have pancreas cancer?* | 100.0% | 100.0% | - | - | - | - | - | - |
| **Overall** | **95.2%** | **96.5%** | **89.6%** | **93.5%** | **96.8%** | **96.2%** | **93.1%** | **94.9%** |

Supplementary Table 3: Performance metrics of the LLM-based classification on the seventeen different questions for the Cetuximab dataset.

| **Question** | **Accuracy** | | **Precision** | | **Recall** | | **F1-score** | |
| --- | --- | --- | --- | --- | --- | --- | --- | --- |
|  | GPT-OSS 20B | GPT-OSS 120B | GPT-OSS 20B | GPT-OSS 120B | GPT-OSS 20B | GPT-OSS 120B | GPT-OSS 20B | GPT-OSS 120B |
| *Is this a Case Report reporting on the history of a single patient?* | 98.0% | 97.0% | 98.6% | 97.3% | 98.6% | 98.6% | 98.6% | 98.0% |
| *Did the patient receive radiation therapy?* | 97.3% | 95.9% | 98.5% | 98.4% | 98.5% | 96.9% | 98.5% | 97.7% |
| *Did the patient receive the systemic therapy Cisplatin?* | 100.0% | 98.6% | 100.0% | 100.0% | 100.0% | 98.2% | 100.0% | 99.1% |
| *Was Cisplatin given before Radiotherapy?* | 85.2% | 88.9% | 69.6% | 76.2% | 94.1% | 94.1% | 80.0% | 84.2% |
| *Was Cisplatin given at the same time as Radiotherapy?* | 90.7% | 83.3% | 96.7% | 96.2% | 87.9% | 75.8% | 92.1% | 84.7% |
| *Was Cisplatin given after RT?* | 96.2% | 98.1% | 93.3% | 100.0% | 93.3% | 93.3% | 93.3% | 96.6% |
| *Did the patient die?* | 100.0% | 100.0% | 100.0% | 100.0% | 100.0% | 100.0% | 100.0% | 100.0% |
| *Did the patient have a positive outcome from the interaction between radiotherapy and the systemic therapy Cisplatin?* | 77.8% | 79.6% | 82.9% | 85.3% | 82.9% | 82.9% | 82.9% | 84.1% |
| *Did the patient have a negative outcome from the interaction between radiotherapy and the systemic therapy Cisplatin?* | 92.6% | 88.9% | 78.6% | 71.4% | 91.7% | 83.3% | 84.6% | 76.9% |
| *Is the patient male?* | 98.1% | 98.1% | 96.2% | 96.2% | 100.0% | 100.0% | 98.0% | 98.0% |
| *Is the patient female?* | 100.0% | 100.0% | 100.0% | 100.0% | 100.0% | 100.0% | 100.0% | 100.0% |
| *Did the patient have lung cancer?* | 98.1% | 100.0% | 90.9% | 100.0% | 100.0% | 100.0% | 95.2% | 100.0% |
| *Did the patient have kidney cancer?* | 100.0% | 100.0% | 100.0% | 100.0% | 100.0% | 100.0% | 100.0% | 100.0% |
| *Did the patient have lymphoma?* | 100.0% | 100.0% | 100.0% | 100.0% | 100.0% | 100.0% | 100.0% | 100.0% |
| *Did the patient have skin cancer?* | 98.1% | 96.3% | 66.7% | 50.0% | 100.0% | 50.0% | 80.0% | 50.0% |
| *Did the patient have a gynecological cancer?* | 100.0% | 100.0% | 100.0% | 100.0% | 100.0% | 100.0% | 100.0% | 100.0% |
| *Did the patient have pancreas cancer?* | 100.0% | 100.0% | 100.0% | 100.0% | 100.0% | 100.0% | 100.0% | 100.0% |
| **Overall** | **96.2%** | **95.7%** | **94.2%** | **94.8%** | **96.1%** | **94.0%** | **95.1%** | **94.4%** |

Supplementary Table 4: Performance metrics of the LLM-based classification on the seventeen different questions for the Cisplatin dataset.
